# Integrative single-cell profiling of melanoma reveals a tumor microenvironment signature predictive of immunotherapy response

**DOI:** 10.64898/2026.05.13.26352980

**Authors:** Theodoros Margelos, Ioanna Mina, Aggeliki Tserga, Eirini Goula, Sophia Kondylis, Antonia Vlahou, Maria Frantzi

## Abstract

**Background:** Immune checkpoint inhibitors have transformed cancer treatment, yet a large number of patients fail to respond. Identifying molecular characteristics that predict response before treatment initiation remains an unmet need. Towards that end, this study presents a large-scale integrative analysis of existing single-cell and bulk tissue datasets, aimed at identifying predictive features while providing insights into their cellular origin and potential function within the tumor microenvironment.

**Methods:** A stepwise analysis was performed using single-cell RNA-sequencing data from 60 melanoma patients at baseline, separated into discovery (n=41) and validation (n=19) sets. An integrated bulk transcriptomics dataset (n=128) from melanoma patients and a bladder cancer dataset (n=298) were used for further validation.

**Results:** Integrative analysis of melanoma single-cell datasets revealed that responders exhibit distinct molecular profiles across multiple cell types compared to non-responders. Notably, these included downregulation of the TNFR superfamily and other immunosuppressive genes (TNFRSF18, TNFRSF9, TNFRSF4, LGALS1, BATF, IL12RB2, LINGO1, DUSP4, SDC4, VCAM1) in T-cells. By investigating the findings from the immune cell populations in the bulk tumor context, 13 transcripts were found to be consistently associated with response across all cohorts. These were differentially expressed in T-cells (SELL, EPB41, CD96, UHFR2, LINGO1, LGALS1), B-cells (ALDH5A1), NK cells (PLEC, PDGFRB) and Monocytes (TLR10, ST6GAL1, IKZF1, MPRIP). A predictive model based on these features effectively discriminated responders from non-responders in melanoma (AUC=0.73). The model maintained significant predictive power in an independent bladder cancer dataset (IMvigor210; AUC=0.64). Of high clinical relevance, it demonstrated enhanced performance in identifying responders among patients with low tumor mutational burden (AUC=0.75).

**Conclusion:** Our study reveals pre-treatment molecular features related to immune-cancer crosstalk that are associated with response to immunotherapy. A 13-gene model demonstrates potential added clinical value in stratifying responders, particularly in patients with low tumor mutational burden, meriting further validation.

## Background

In recent years, immunotherapy particularly in the form of immune checkpoint inhibitors (ICI) targeting PD-1, CTLA4 and LAG3, has become an integral component in the treatment of advanced tumors. High-risk melanoma and advanced urothelial cancer represent key indicators, with evidence demonstrating improved survival outcomes after treatment [1][2]. However, only approximately 33-58% of melanoma patients receiving immunotherapy respond to ICIs [3][4,5], while the respective overall response rate (ORR) in bladder cancer remains even lower, ranging from 17% to 29% [6,7]. Although the combination of ICI regimens can improve ORR, it is often related to increased adverse events [8].

To better stratify patients, several predictive biomarkers have been proposed, including tumor mutational burden (TMB) and PD-L1 expression assessed by immunohistochemistry. High TMB has been associated with improved response to ICIs, likely reflecting an increased neoantigen load that enhances tumor immunogenicity [9]. However, its predictive value remains inconsistent across tumor types and individual patients, limiting its clinical utility as a standalone biomarker [10,11]. Similarly, PD-L1 expression has been widely used to guide treatment decisions, yet it suffers from significant limitations, including staining variability even within the same tumor (intratumoral heterogeneity), alongside differences in assay platforms and scoring thresholds, all of which contribute to poor reproducibility and limited predictive efficacy [12,13].

Most non-responders (NRs) exhibit primary resistance [14], indicating that immunosuppressive mechanisms are often present prior to treatment initiation, rather than acquired during its course. Consequently, the patient’s pre-treatment (baseline) molecular profile may hold significant predictive value [15]. Resistance may stem from pre-existing immune system dysfunction and/or tumor intrinsic characteristics within the tumor microenvironment (TME) [16], including immune-desert phenotypes characterized by low T-cell generation, as well as dysfunctional or exhausted T-cell states that impair effective anti-tumor immunity [17]. Transcriptomics [18–21] and proteomics studies [22–24] have identified molecular features associated with immunotherapy response and resistance in melanoma patients. For instance, interferon-γ (IFN-γ) signaling correlates with enhanced antitumor immunity [25] and activation of STAT1 and Wnt-pathway genes has been linked to favorable outcomes [19][26]. On the contrary, upregulation of inhibitory receptors, such as TIM-3 was associated with resistance [24]. Similarly, in the case of urothelial carcinoma T-cell inflammatory and interferon-related gene signatures, as well as IFN-γ expression, have demonstrated predictive value for response to ICIs [16].

These existing studies, even though informative, often lack detailed insights at the cellular level. This limitation is addressed by the implementation of single-cell technologies, which enable the characterization of cell-type specific molecular alterations associated with immunotherapy resistance [27]. Prior work has reported that reduced tumor-immune cell interactions and increased abundance of CD4+ CXCL13+ follicular helper-like T-cells in the TME are linked to ICI resistance [28,29]. To date, only a limited number of meta-analyses have investigated single-cell datasets, in the context of immunotherapy response, primarily focusing on during or post-treatment samples in both melanoma-specific and pan-cancer contexts [30,31]. Nevertheless, no comprehensive analysis has investigated pre-treatment single-cell datasets, despite their potential to identify cell-type specific features associating with treatment response and to elucidate their cellular origin and potential underlying biological mechanisms depicting tumor-TME crosstalk. To address this gap, in this study, we performed a large-scale integrative analysis of pre-treatment single-cell transcriptomics datasets, followed by validation in bulk tissue to increase translatability, targeting the identification of predictive molecular features across diverse patient populations, clinical settings, and tumor entities treated with ICIs. Our results highlight common immune modules associated with responsiveness, that may provide added predictive value over current molecular prognosticators.

## Methods

### Single-cell RNA sequencing data retrieval

To identify single-cell RNA-sequencing (scRNA-seq) datasets relevant to melanoma immunotherapy response, a literature search was conducted using the keywords melanoma (AND) single-cell RNA (AND) human in Medline. Inclusion criteria involved (1) treatment with ICI, specifically antibodies targeting PD-1, PD-L1, CTLA-4, LAG-3 or their combination, (2) availability of baseline (pre-treatment) transcriptomic data (3) reported immunotherapy response outcomes (4) clear links (identifiers) of the clinical with molecular data. Studies involving blood mononuclear cell (PBMCs) samples, uveal melanoma, cell lines, animal models, only post-treatment data, or review articles were excluded. Three datasets met the inclusion criteria: two with raw count data (E-MTAB-13770 [32], EGAS00001006488 [33] and one with transcripts per million (TPM) values [34]. The first two were integrated into a discovery cohort while the TPM dataset was used as an initial single-cell validation cohort.

### Single-cell RNA sequencing data processing

ScRNA-seq data were processed and analyzed using the Seurat package in R [35]. To ensure data quality, filtering criteria were applied to remove low-quality cells and potential doublets. Cells were retained if they presented counts in 500 to 5.000 detected genes. To eliminate dying cells, cells with mitochondrial gene content exceeding 20% were excluded. To omit erythrocytes from the dataset, cells with more than 5% of hemoglobin gene content were also removed. Raw gene expression counts were normalized using the *NormalizeData* function in Seurat, applying log-normalization with a scale factor of 10. The 3,000 most variable genes across cells were identified using the *FindVariableFeatures* function with the variance-stabilizing transformation method. The normalized data were then scaled using the *ScaleData* function. Principal component analysis (PCA) was performed using the *RunPCA* function to calculate the top 50 principal components (PCs) based on the 3,000 variable genes. To mitigate technical batch effects arising from sample-specific and study-specific differences, datasets within the discovery cohort were integrated using the *RunHarmony* function from the Harmony package. The same function but only for sample-specific differences was used in the validation cohort. Cell clustering was conducted by constructing a shared nearest-neighbor (SNN) graph using the *FindNeighbors* function, based on the first 20 PCs from the Harmony-integrated dataset. Clusters were identified using the *FindClusters* function with the Louvain algorithm at a resolution of 0.5. Uniform Manifold Approximation and Projection (UMAP) was performed using the *RunUMAP* function on the integrated PCs to visualize cell clusters in two-dimensional space.

### Cell type annotation

In the discovery cohort, clusters expressing marker genes for melanoma cells, endothelial cells, and fibroblasts as defined by Tirosh et al. [36] were annotated accordingly. The remaining cells of the discovery cohort were all positive for protein tyrosine phosphatase, receptor type C (PTPRC or CD45+) indicating that they belong to immune cell populations (**Supplementary Figure 1**.). In the validation set, all cells were derived from immune populations (CD45+). These immune cells from both discovery and validation cohorts were annotated using the SingleR package [37] utilizing the Blueprint Encode database from R package celldex (1.18.0) as reference [38]. Due to the low number of cells identified as Dendritic Cells, Macrophages, and Neutrophils (fewer than 55 cells per population in the validation cohort), these lineages were annotated as Monocytes. This reassignment ensured sufficient cellular depth per patient for reliable pseudobulk aggregation. Marker genes for each cell type were identified using the Seurat *FindAllMarkers* function, with its default parameters. To annotate the functional states of T-cells in the discovery cohort the R package Project TILS was implemented [39]. Specifically, the previously annotated CD4+ and CD8+ T-cells were projected in the included subtypes, using the CD4+ and CD8+ human reference maps of the package. This process was followed only in the discovery cohort, due to the small number of T-cells in the validation cohort.

### Single-cell differential expression analysis

To evaluate differential gene expression between Rs and NRs within each cell type, a pseudobulk approach was employed. This approach treats the individual patient, rather than the single-cell, as the unit of independent observation, thereby preventing the inflation of false-positive rates. Specifically, raw UMI counts were aggregated by summing gene expression across all cells per patient and cell type using the AggregateExpression function in Seurat. These pseudobulk expression matrices from the discovery cohort were then analyzed using the DESeq2 package in R [40], where library size normalization occurs. To account for technical variability across datasets, the ‘Study’ variable was included as a covariate in the design formula.

For the validation cohort, differential expression was assessed on aggregated patient-level TPM data using the limma R package [41]. To meet the assumptions of linear modeling, TPM values were log-transformed prior to analysis. We employed sample-specific array weights to account for varying sample quality and cell-count-derived noise, and additionally the analysis was restricted to cell types with at least three biological replicates per group. Models were moderated via empirical Bayes with the trend=TRUE parameter to capture the mean-variance relationship of the log-transformed data.

### Bulk RNA sequencing data retrieval and analysis

Four publicly available RNA-seq datasets [42–45], containing baseline samples from melanoma patients prior to immunotherapy, along with available response outcomes, were retrieved following a systematic search in Medline using similar search criteria as in the case of the single-cell datasets. The selected datasets were retrieved from the GEO database (GSE78220 [44], GSE213145 [42]), GitHub [43] and Array-Express (E-MTAB-11729) [45]. Data corresponding to the baseline samples from each study were integrated into one RNA-seq validation cohort, by keeping the 11,663 common genes amongst them. Batch effects were corrected in R using the ComBat_seq function from the sva package, with study specified as the batch factor [46]. In order to preserve biologic variability associated with treatment response, response was included in the ComBat_seq model as the biological condition of interest. Following batch correction, the DESeq2 R package was used to normalize the data and identify differentially expressed genes between Rs and NRs.

For external validation of the model performance, RNA-seq data and clinical information from pretreatment tumor samples of urothelial carcinoma patients treated with anti-PD-L1 therapy were obtained from the IMvigor210 phase II clinical trial using the IMvigor210CoreBiologies R package [47]. The urothelial carcinoma dataset, containing information about the expression of 24,033 genes, was normalized following the same normalization process used to normalize the bulk RNA melanoma data using the DESeq2 package. To evaluate whether there was a consistent technical variation between the melanoma and bladder cancer bulk RNA data, only their common 11,645 genes were selected. The median gene expression levels across these genes in the bladder cancer dataset was 5.3 times higher than in the melanoma dataset and as such, the DESeq2 expression values of the former were divided by 5.3 to normalize the bladder cancer relative to the melanoma dataset.

### Model development

The logistic regression model was trained to predict response to ICI using the caret package in R. Model training and internal validation were performed in the normalized melanoma bulk RNA-seq dataset, using 10-fold cross-validation. Predicted probabilities from all the cross-validation folds were combined to generate a cross-validated receiver operating characteristic (ROC) curve. A probability threshold corresponding to approximately 90% sensitivity was determined from these pooled cross-validated predictions. This threshold was then applied to the external normalized urothelial cancer bulk RNA-seq dataset. To further evaluate model performance in tumors with low tumor mutational burden (TMB), the model was evaluated in the subset of patients with TMB <10 mutations per Mb (mut/Mb), a commonly used clinical threshold for defining low-TMB tumors [48]. To plot the ROC curves, calculate the area under the curve (AUC) with 95% confidence intervals and estimate sensitivity and specificity at specific thresholds the pROC package in R was used. To assess the statistical significance of the AUC, the roc.area function from the verification package in R was used.

The resulting scores (linear predictor) of the model for each patient were then correlated with the whole transcriptome expression of the 2 datasets using the corr.test function from the psych R package [49]. Genes exhibiting a positive or negative spearman correlation with an absolute |r| > 0.3 were selected for pathway analysis based on the Gene Ontology Biological Processes (GO:BP) database using the clusterProfiler R package [44]. Overrepresentation analysis (ORA) was conducted for four distinct gene sets (positively and negatively correlated genes to the signature score in each one of melanoma and bladder datasets).

## Results

### Stepwise discovery and validation study design

To identify robust features associated with ICI response a stepwise discovery and validation analysis was conducted (workflow shown in **Figure 1**). Based on data accessibility, a discovery dataset consisting of two independent melanoma scRNA-seq datasets [32,33] was generated. In brief, the discovery dataset included 42 baseline samples (n= 23 from Pozniak et al. [33], n= 19 from Schlenker et al. [32]) originating from 41 patients. Findings from this dataset were then validated initially in an independent melanoma validation scRNA-seq dataset [34]. In the discovery cohort a total of 106,837 cells were included, comprising CD8+ T-cells (35.78%), CD4+ T-cells (16.19%), B-cells (15.35%), monocytes (12.39%), melanoma cells (8.32%), NK cells (4.20%), macrophages (3.91%), fibroblasts (2.69%), dendritic cells (0.66%), and endothelial cells (0.49%), corresponding to 88.5% immune and 11.5% non-immune cell types (**Figure 2A**). The validation dataset included 5,905 cells, most of which were immune cells (98.99%), comprising CD8+ T-cells (50.38%), CD4+ T-cells (23.52%), B-cells (11.53%), monocytes (8.04%), NK cells (5.50%), and a small proportion of hematopoietic stem cells (HSCs, 1.02%) (**Figure 2B**). Cell type identities were validated and supported by distinct marker gene expression profiles, as illustrated by the top differentially expressed genes per cell type (**Supplementary Figure 2**).

**Figure 1.**
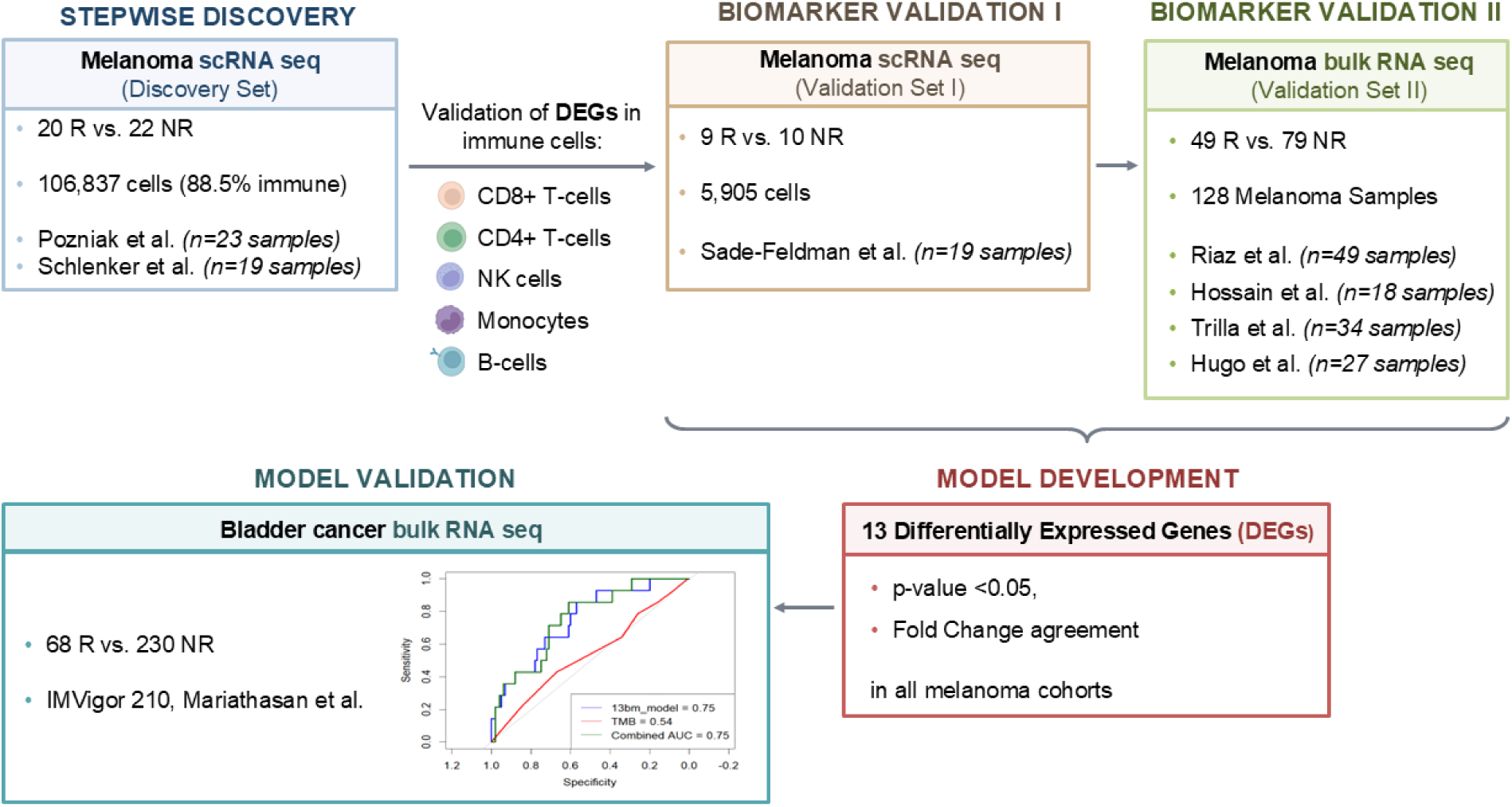
Study workflow. Integrative analysis of curated transcriptomics datasets was conducted to identify gene expression patterns associated with immunotherapy response. Candidate differentially expressed genes (DEGs) were first identified in a melanoma single-cell RNA sequencing (scRNA-seq) discovery cohort (n=42) across five immune cell lineages (CD4+/CD8+ T-cells, NK cells, Monocytes, B-cells) and were replicated in an independent single-cell dataset (n=19). DEGs which were subsequently validated in an integrated bulk RNA-seq (n=128) melanoma cohort were selected for model development. Specifically, a 13-gene signature was used to train a predictive model which was further tested in an external bladder cancer cohort (n=298).

**Figure 2.**
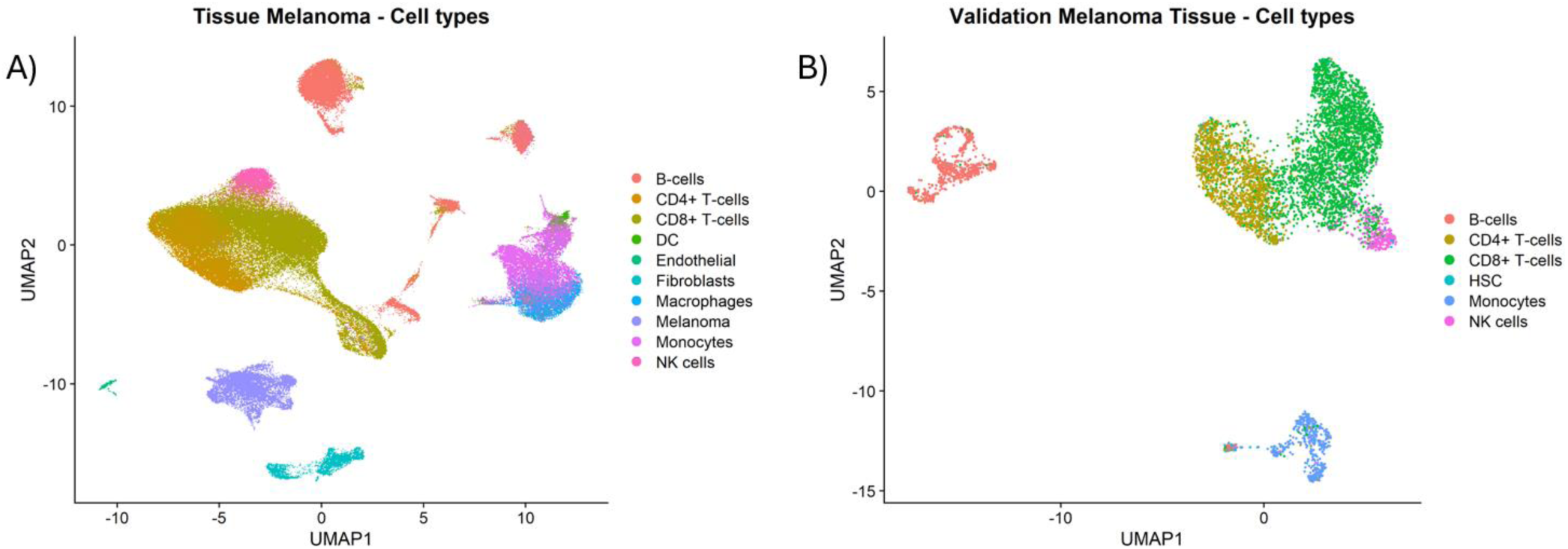
Cellular composition of the 2 single-cell cohorts. UMAP visualizations showing the cellular composition of the (A) single-cell discovery cohort and (B) the single-cell validation cohort. UMAP: Uniform Manifold Approximation and Projection

The validated scRNA-seq findings were further tested for their detection and differential expression within melanoma bulk RNA-seq data similarly collected prior to ICI treatment and with available treatment response outcomes. The melanoma bulk RNA-seq cohort included 128 samples from 4 different studies: 49 samples from Riaz et al. [43], 18 samples from Hossain et al. [42], 34 samples from Trilla et al. [45] and 27 samples from Hugo et al. [44]. Shared differential expression genes (DEGs) across the melanoma single-cell and bulk cohorts were then used to construct a predictive model. To evaluate the clinical utility and potential pan-cancer applicability of this model, an additional validation in an independent bladder cancer bulk RNA-seq cohort (IMvigor210, n = 298) [47] was further performed.

All samples used in this study were collected prior to the induction of the ICI - treatment. Clinical and demographic information for each cohort, including treatment scheme and disease stage at inclusion, is presented in **Table 1**. In the melanoma single-cell discovery and bulk cohorts and in the bladder cancer cohort response was assessed as Complete Remission (CR), Partial Remission (PR) or Tumor Free (TF). Non-response was evaluated as Stable (SD) or Progressive disease (PD). In the melanoma single-cell validation cohort, response was based on radiologic assessments and classified as progression/NRs or regression/Rs. To ensure consistency across the bulk transcriptomics datasets, response categories were harmonized such that CR and PR were grouped as Rs, and SD and PD as NRs.

**Table 1.** Clinical Summary of the datasets used in the analysis. Clinical cohorts included in the study with relevant treatment information and basic demographics, as per availability. PD-1: anti PD-1 monotherapy, CTLA-4: anti-CTLA-4 monotherapy, PD-L1: anti-PDL1 monotherapy, PD-1 + LAG-3: combination anti-PD1 and anti-LAG3 therapy, PD-1 + CTLA-4: combination anti-PD1 and anti-LAG3 therapy. F: Female, M: Male, *NA* (Not Available) indicates clinical data not provided in the original data repositories

| Cohort | Total Samples | Responders (%) | Non-Responders (%) | Treatment Breakdown | Stages | Gender | Age |
| --- | --- | --- | --- | --- | --- | --- | --- |
| Tissue Discovery scRNA [32,33] | 42 | 22 (52.3%) | 20 (47.7%) | PD-1: 26 (62%)<br>PD-1 + CTLA-4: 13 (31%)<br>CTLA-4: 2 (4.7%)<br>PD-1 + LAG3: 1 (2.4%) | M0: 17 (40.5%),<br>M1: 25 (59.5%) | F: 20 (47.6%),<br>M: 22 (52.4%) | ≥60: 25 (59.5%),<br><60: 17 (40.5%) |
| Validation scRNA [34] | 19 | 9 (47.4%) | 10 (52.6%) | PD-1: 12 (63.2%)<br>PD-1 + CTLA-4: 5 (26.3%)<br>CTLA-4: 2 (10.5%) | M1 (100%) | F: 6 (31.6%),<br>M: 13 (68.4%) | ≥60: 15 (78.9%),<br><60: 4 (21.1%) |
| Melanoma Bulk Transcriptomics [42–45] | 128 | 49 (38.3%) | 79 (61.7%) | PD-1: 128 (100%) | M0: 2 (1.6%), M1: 66 (51.6%),<br>NA: 60 (46.8%) | F: 13 (10.2%),<br>M: 48 (37.5%),<br>NA: 67 (52.3%) | ≥60: 40 (31.3%),<br><60: 21 (16.4%),<br>NA: 67 (52.3%) |
| Bladder Bulk Transcriptomics [50] | 298 | 68 (22.8%) | 230 (77.18%) | PD-L1: 298 (100%) | M1: (100%) | F: 65 (21.8%),<br>M: 233 (78.2%) | NA |

### Molecular profiles of tumor infiltrating immune cells associating with immunotherapy response

Differential expression analysis (DEA) of pseudo-bulk aggregated counts from specific cell types was conducted to identify features significantly and consistently associated with response to ICIs (DESeq2 nominal p-value < 0.05), both in the discovery (**Supplementary Table 1)** and the validation (**Supplementary Table 2**) cohorts. In the tumor infiltrating CD4+ T-cells, 37 transcripts (9 upregulated, 28 downregulated in Rs) were found to be commonly differentially expressed in Rs versus NRs in both single-cell discovery and single-cell validation sets (**Supplementary Table 3-CD4+ cells**), whereas the respective number for CD8+ T-cells was 108 genes (35 upregulated, 73 downregulated in Rs versus NRs) (**Supplementary Table 3-CD8+ cells**).

Interestingly, in both cell types, a consistent prominent downregulation of the tumor necrosis factor receptor superfamily genes TNFRSF18 (GITR) and TNFRSF9 (4-1BB) was observed in Rs, while TNFRSF4 (OX40) was also downregulated in CD8+ T-cells of Rs versus NRs (**Table 2**). When investigating the expression profiles of the TNFRSF genes in distinct T-cell functional states (**Figure 3A**), TNFRSF9 displayed elevated expression levels across both the CD8+ exhausted and CD8+ progenitor exhausted lineages, while TNFRSF18 and TNFRSF4 presented peak expression within the CD4+ regulatory and CD4+ cytotoxic exhausted clusters (**Figure 3B**). This distribution of expression in regulatory and exhausted T-cell lineages was also observed for additional genes implicated in immune cell regulation (**Table 2**, **Figure 3B, Supplementary Table 4**).

**Figure 3.**
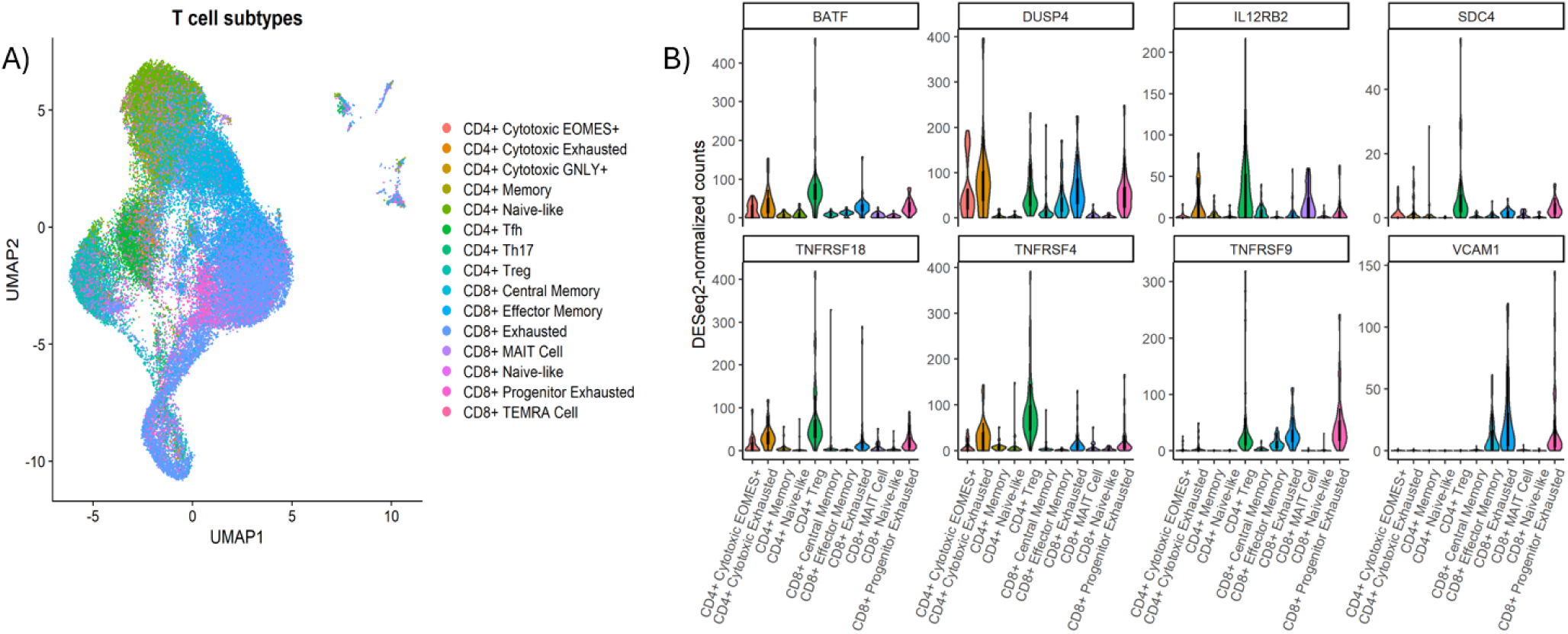
Transcriptomic Profiling and Marker Gene Expression of T-Cell Subsets. (A) UMAP visualization of T-cells from the single-cell discovery cohort annotated into their functional states using the ProjectTILs reference atlas. (B) Violin plots showing DESeq2-normalized expression of key genes (TNFRSF4, TNFRSF9, TNFRSF18, IL12RB2, SDC4, VCAM1, DUSP4, BATF) across T-cell subtypes, stratified by patient response. UMAP: Uniform Manifold Approximation and Projection, CD4+ Treg: CD4+ Regulatory T-cells, CD4+ Tfh: CD4+ Follicular Helper T-cells, CD4+ Th17: CD4+ Helper T-cells type 17, CD8+ TEMRA Cell: CD8+ Terminally Differentiated Effector Memory T-cells re-expressing CD45RA, CD8+ MAIT Cell: CD8+ Mucosal-Associated Invariant T-cells.

**Table 2.**
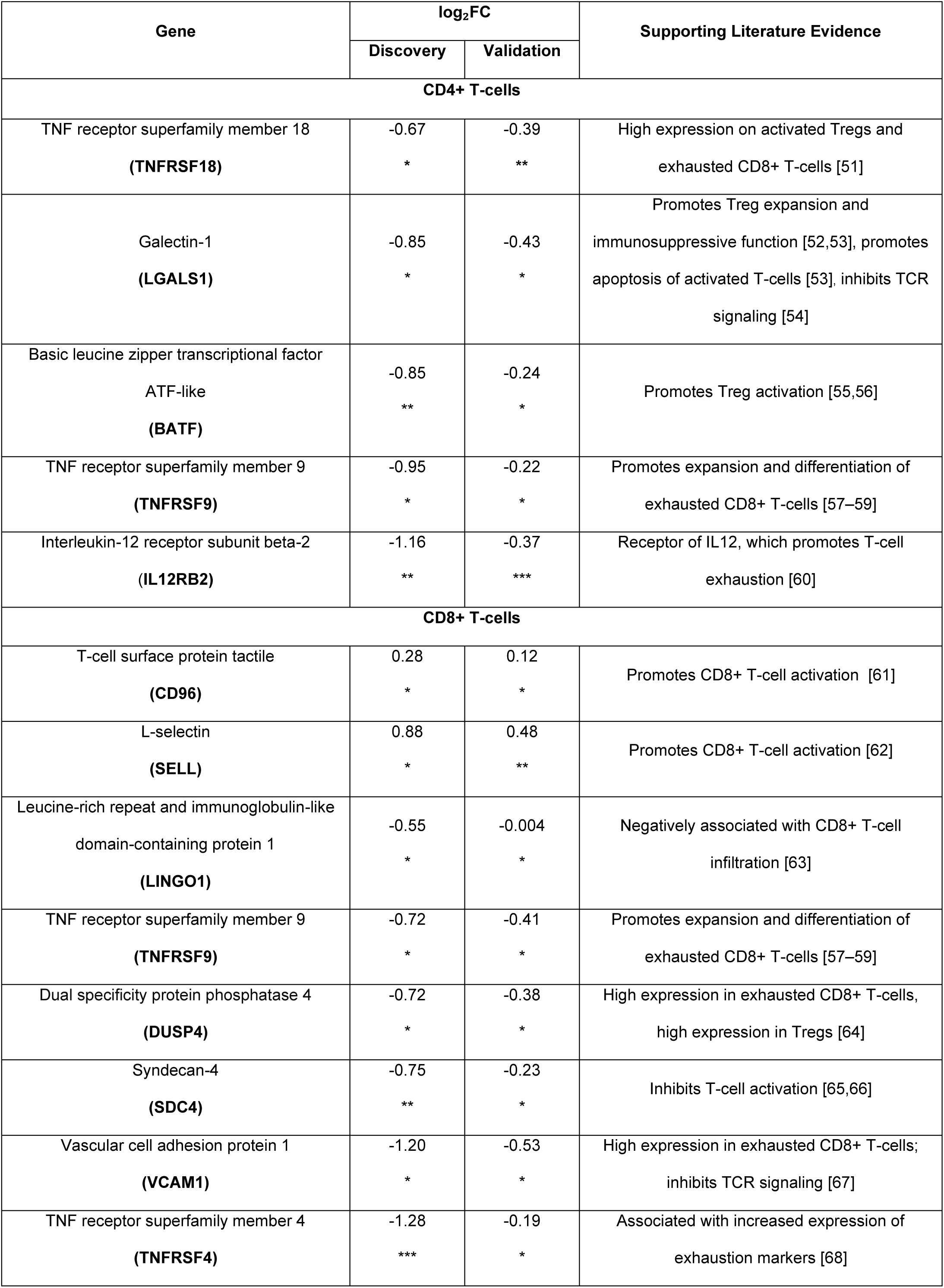
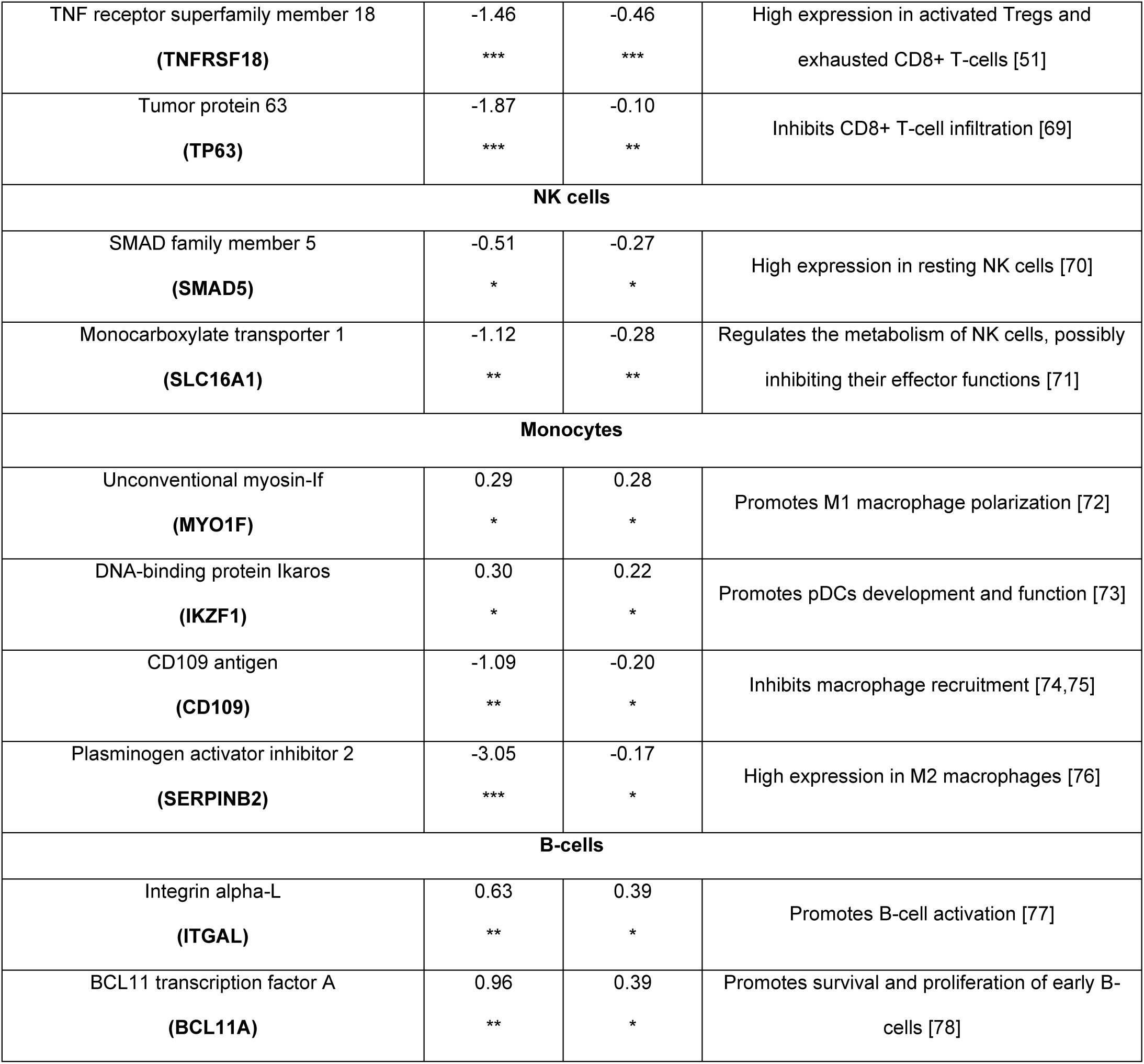
Differential expression of Immunotherapy response related genes across discovery and validation cohorts. Representative genes associated with response to ICIs (DESeq2 nominal p-value < 0.05) in both discovery and validation scRNA datasets, with known roles in immune cell regulation. Log₂ fold-change (log₂FC) values from DESeq2 pseudo-bulk differential expression analysis are shown for each cohort. Positive log₂FC values indicate higher expression in Rs, whereas negative values indicate lower expression in Rs relative to NRs. Statistical significance based on DESeq2 nominal p-value: *p-value < 0.05, **p-value < 0.01, ***p-value < 0.001. IL12: Interleukin-12, pDCs: plasmacytoid dendritic cells, Tregs: regulatory T-cells, TCR (T-cell Receptor)

Regarding the rest cell types, in tumor-infiltrating NK cells, 48 transcripts were downregulated in Rs versus NRs across both the discovery and validation datasets (**Supplementary Table 3 - NK cells**), while 48 transcripts (22 upregulated, 26 downregulated in Rs versus NRs) were consistently differentially expressed in tumor infiltrating monocytes of both discovery and validation sets (**Supplementary Table 3 - Monocytes**). In B-cells, 31 transcripts were found to be commonly differentially expressed in Rs versus NRs in both discovery and validation sets (17 upregulated, 14 downregulated) (**Supplementary Table 3 - B cells**). In all cases, several of the differentially expressed genes had been reported in the literature for their functional relevance to immune cell regulation, as summarized in **Table 2**, further supporting the validity of our stepwise applied approach.

### A 13-gene signature predicts response to ICI

To get an insight into the translational potential of these observations, their investigation at the bulk tissue level was performed. Thirteen genes were found to be significantly associated with response to ICI (DESeq2 nominal p-value < 0.05) in the melanoma single-cell discovery and validation cohorts and in the melanoma bulk cohort (**Supplementary Table 5**), while maintaining the same regulation trend across all three cohorts (**Table 3**).

**Table 3.**
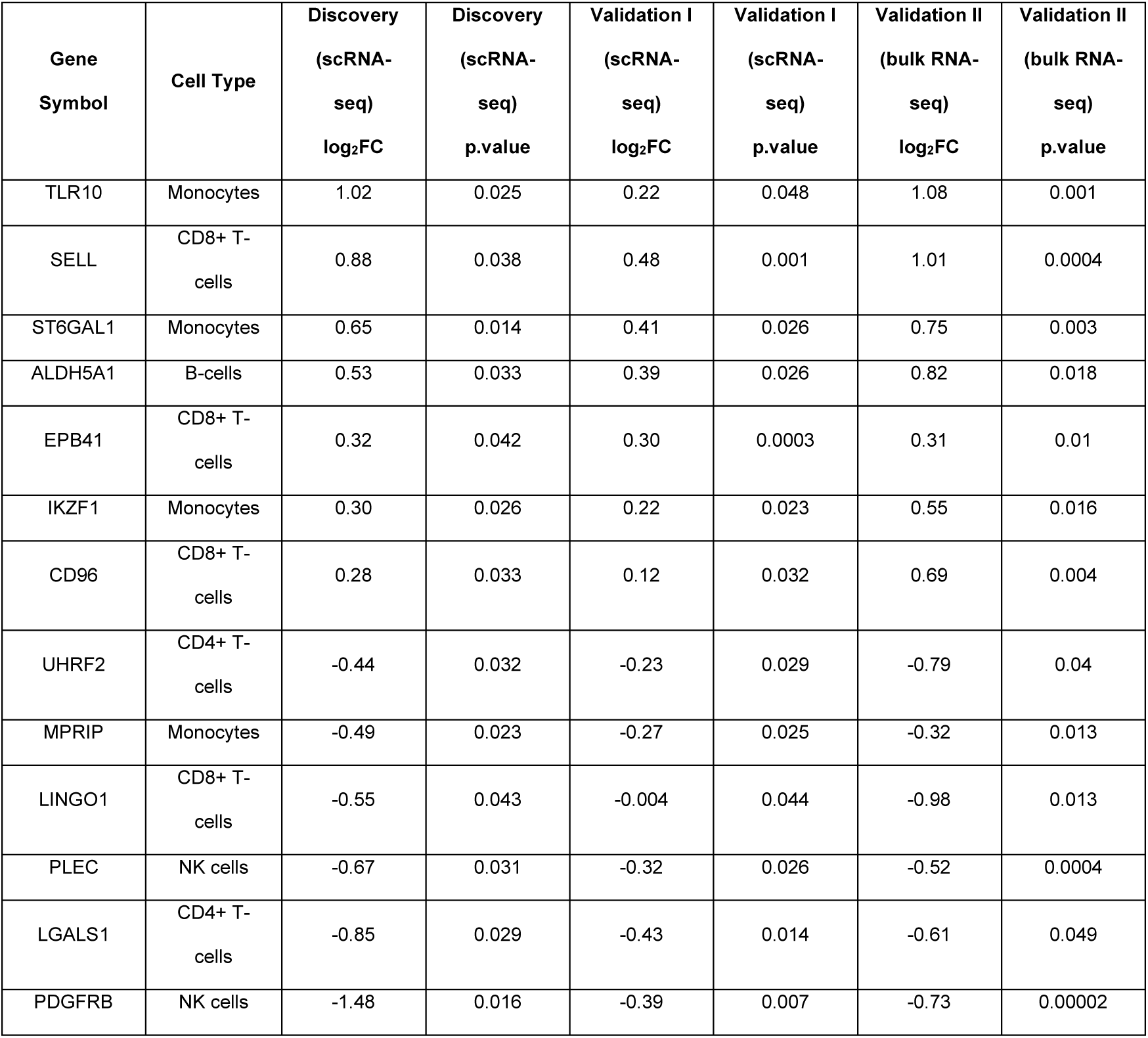
Transcripts whose levels are consistently associated with response to immune checkpoint inhibitors (ICI) in melanoma. Expression was investigated across melanoma single-cell (discovery and validation I) and bulk RNA-seq cohorts (Validation II). The presented 13 genes were significantly associated with response to ICI (DESeq2 nominal p-value < 0.05) and exhibited consistent fold change (FC) trend in all cohorts. log₂FC: log₂-transformed fold change in expression between responders (Rs) and non-responders (NRs) estimated with DESeq2, with positive values indicating higher expression in Rs and negative values indicating higher expression in NRs.

A logistic regression model based on the 13 biomarkers (13BM) achieved an AUC of 0.73 (95% CI 0.65-0.82, p=5.1*10⁻⁶) in the melanoma bulk RNA-seq cohort, based on 10-fold cross-validation (**Figure 4A**). At the default probability threshold of p_r_ = 0.5, sensitivity for detection of Rs was 49.0% and specificity was 78.5%. Since increased sensitivity in the identification of responders to ICI would be of added clinical value, a lower probability threshold (p_r_ = 0.19) was selected, corresponding to approximately 90% sensitivity. Using this threshold, the model achieved a sensitivity of 89.8% and a specificity of 40.5% in cross-validation.

**Figure 4.**
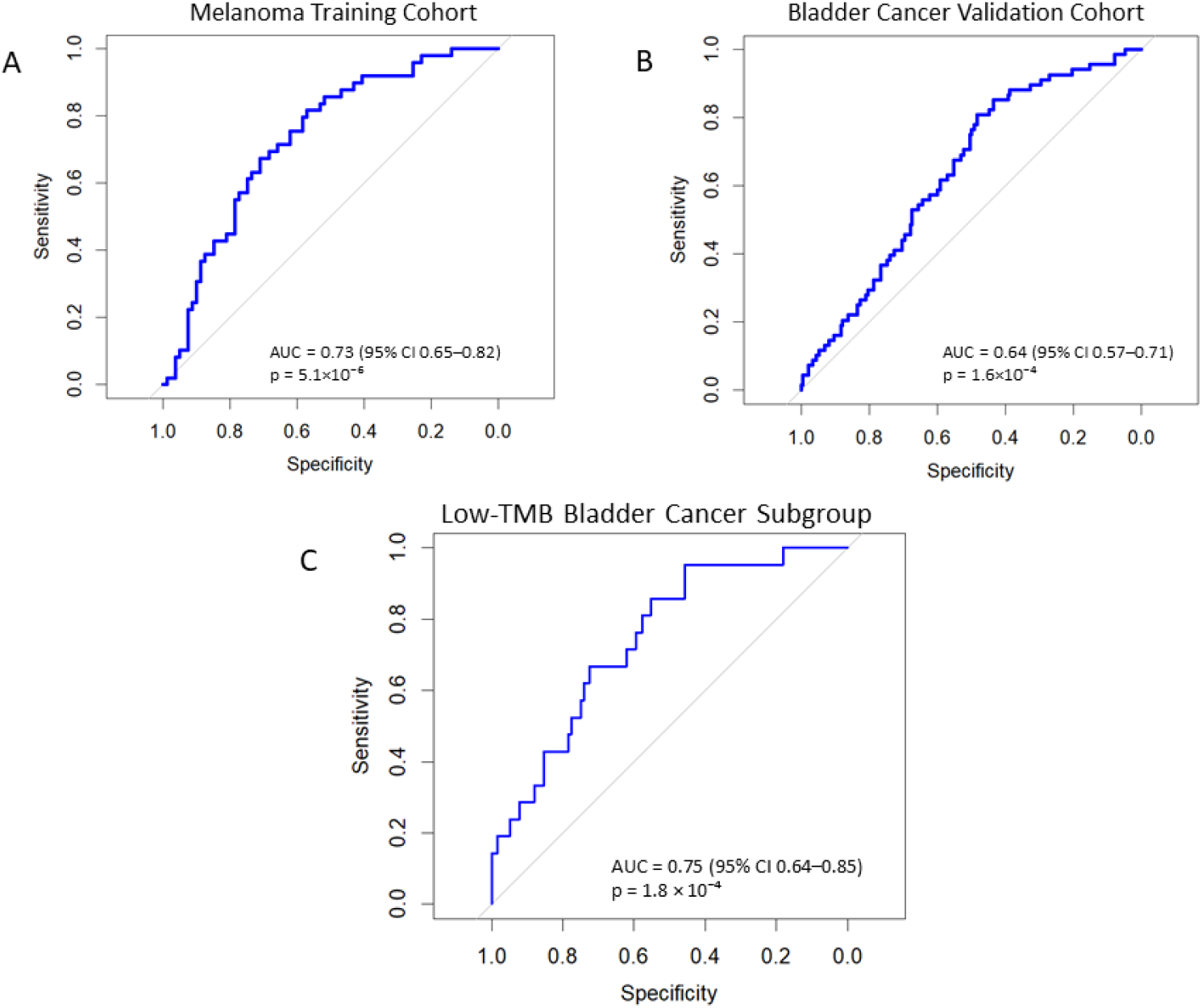
Performance of the 13BM in predicting response to immune checkpoint blockade. (A) Receiver operating characteristic (ROC) curve in the melanoma bulk RNA-seq cohort using 10-fold cross-validation. (B) ROC curve in the independent bladder cancer bulk RNA-seq validation cohort. (C) ROC curve in the low-TMB subgroup defined as less than 10 mutations per Mb (n = 137, 21 Rs). 13BM: 13 Biomarkers Model, TMB: Tumor Mutational Burden

**Figure 5.**
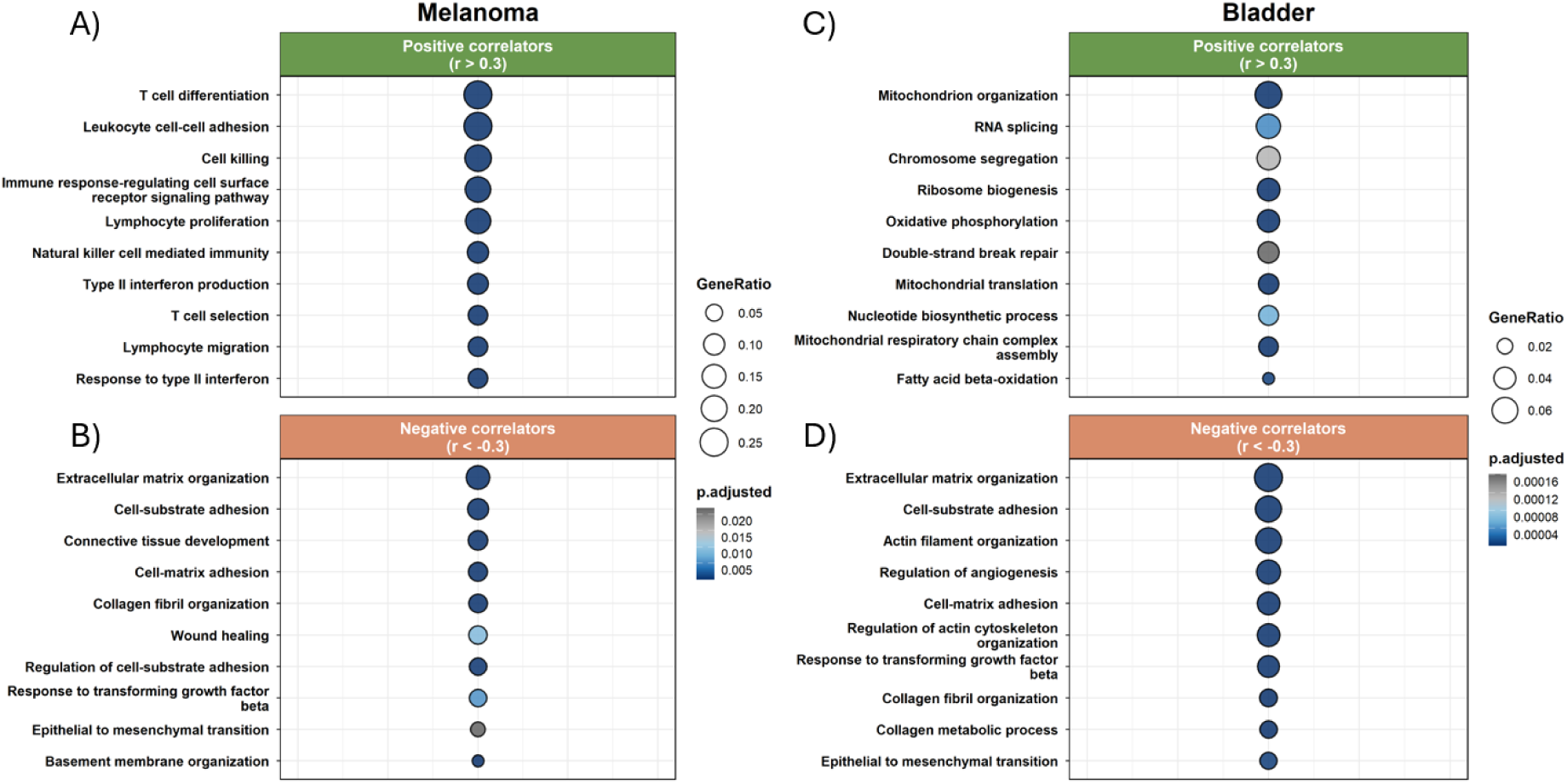
Gene set overrepresentation analysis (ORA) of genes correlating with the 13BM signature score. The dot plots show Gene Ontology (GO) Biological Processes enriched in genes positively (Spearman r > 0.3) or negatively (Spearman r <0.3) correlated with the 13BM score; (A-B) the bulk melanoma and (C-D) bladder cancer cohorts.

Interestingly, when applied in the bladder cancer bulk RNA validation cohort, the model maintained an AUC of 0.64 (95% CI 0.57-0.71, p =1.6*10⁻⁴) **(Figure 4B)**. Applying the same predefined threshold (p_r_ = 0.19), the model discriminated Rs from NRs with 88.2% sensitivity and 38.3% specificity, similarly to the melanoma cohort.

Given that information on TMB was available for the bladder cancer cohort, a further investigation was performed to test the complementarity of the two molecular prognosticators (TMB and 13BM). A threshold of ≥10 mut/Mb commonly used to define TMB-high tumours [79] was applied; as such, a group of 137 patients (including 21 Rs) with TMB <10 mut/Mb was generated (theoretically excluded from ICI treatment based on the TMB prognosticator [79]). Interestingly, in this subset, the model achieved an AUC of 0.75 (95% CI 0.64–0.85, p=1.8*10^-4^) (**Figure 4C**). Based on the predefined threshold (p_r_ = 0.19), the model achieved 95.2% sensitivity and 39.7% specificity in this cohort, discriminating 20 of 21 Rs who would have otherwise been excluded from treatment.

To gain more insights into the TME biology reflected by 13BM, a correlation analysis of the calculated 13BM score with the global transcriptome of each patient, followed by functional and pathway annotation were performed **(Supplementary Table 6)**. In the melanoma cohort, 13BM correlated positively mainly with transcripts segregating into immune-related pathways, particularly gene sets involved in T-cell differentiation and selection, leukocyte adhesion, proliferation or killer function, interferon production and response, suggesting that the signature reflects an ‘immune-hot’ TME. Surprisingly, in the bladder cancer cohort, 13BM had strongest positive correlations with genes reflecting mitochondrial function, oxidative phosphorylation and metabolic processes (**Figure 4C**).

On the other hand, interestingly, in both melanoma and bladder cancer, 13BM negatively correlated with transcripts reflecting extracellular matrix (ECM) and its organization (**Figure 4B, 4D**), including epithelial-to-mesenchymal transition (EMT), and response to transforming growth factor-beta (TGF-b).

## Discussion

Although ICI therapies have significantly improved clinical outcomes, a substantial proportion of patients do not respond to treatment. This resistance may arise from pre-existing immune dysfunctions and intratumor features of the TME [16]. Consequently, defining the molecular and cellular determinants of resistance prior to treatment is critical for the improvement of patient stratification. In this study, we analyzed gene expression of immune cell populations from pre-treatment ICI melanoma single-cell datasets, to characterize features underlying response and resistance at high power.

Intriguingly, we observed decreased expression of TNFRSF superfamily genes in T-cells, including TNFRSF9 (4-1BB), TNFRSF4 (OX40), and TNFRSF18 (GITR) in Rs compared to NRs. Notably, TNFRSF4 which was downregulated in CD8+ T-cells of Rs has been linked to poor prognosis in non-small-cell lung cancer and hepatocellular carcinoma (HCC), and its high expression in HCC is linked with enrichment of exhaustion-associated transcriptional programs [68,80]. Moreover, experimental TNFRSF4 stimulation was found to antagonize the effects of PD-1 blockade by inducing T-cell apoptosis [81]. In addition, consistently, both CD4+ and CD8+ T-cells from Rs showed reduced expression of TNFRSF9 and TNFRSF18. TNFRSF9 expressing CD8+ T-cells have been reported to express high levels of exhaustion markers [58,59] and their intratumoral abundance correlates with poorer overall survival (OS) in renal cell carcinoma datasets [82]. Moreover, TNFRSF9 stimulation has been shown to drive terminal differentiation and exhaustion of CD8+ T-cells in murine models [57]. Similarly, TNFRSF18 expression in intratumoral CD4+ T-cells has been associated with poor prognosis in gastric cancer [83], and its expression in CD8+ tumor-infiltrating lymphocytes been linked to exhaustion signatures in colorectal cancer [51,84]. The upregulation of these coreceptor genes in immunosuppressive T-cell functional states, as supported by our analysis, such as in the T-regulatory cells and in the exhausted and progenitor exhausted T-cells is also reflective of their potential role in immune evasion; although we have to note that significant differences in the number of these T-cell states between Rs vs NRs were not observed.

Collectively, these findings coupled with the above-mentioned scientific literature suggest that excessive stimulation of these TNFRSF members may reinforce T-cell dysfunction rather than promote productive effector activation. Of note, many drug agonists that stimulate these receptors have been developed, in order to enhance T-cell activity and evoke antitumor immunosurveillance. However, these efforts had limited efficacy in clinical trials [85–89]. One possible explanation for the latter, based on our results, is that these agonists stimulate regulatory and exhausted T-cell populations, likely promoting their dysfunctional activation rather than their reinvigoration.

Other than the TNFRSF members, T-cells from Rs were characterized by downregulation of additional genes potentially linked to T-cell regulatory functions or exhaustion (LGALS1, BATF, IL12RB2, VCAM1, DUSP4) [52–56,60,64,67]. Such altered expression patterns are also presented on monocytes, with MYOF1 which associates with differentiation to M1 macrophages [72] being higher in Rs; while SERPINB2 which conversely is correlated with differentiation to M2 macrophages [76] being lower in Rs. Additionally, genes regulating immune cell infiltration (TP63, CD109) were also differentially expressed between Rs and NRs [69,75].

The translation of the single-cell findings to the bulk tissue, where clinical application is more feasible, was further investigated using a dataset integrating melanoma bulk RNA-seq datasets, as per availability. The transition from single-cell to bulk tissue poses challenges due to the cellular admixture and dilution of cell type-specific signals. Along these lines, the abovementioned changes in the TNFSFR genes could only be observed at the single-cell level. Nevertheless, and despite this challenge, 13 transcripts preserved their differential abundance levels across both melanoma single-cell and the integrated bulk tissue datasets and were thus prioritized for further analysis. As expected, these markers capture immune cell–derived transcriptional information but their levels also reflect contributions from surrounding stromal and malignant cells.

Several of the 13 prioritized features have been previously linked to disease prognosis and response to ICI, supporting their biological and clinical relevance. For example, the PD-L1 transcription factor IKZF1, which was upregulated in Rs, has been associated with longer OS and response to ICI in osteosarcoma and sarcoma patients respectively [90,91]. Similarly, increased expression of ALDH5A1, which was also upregulated in Rs, is associated with improved OS in ovarian [92,93] and pancreatic tumors [94].

Conversely, features from the 13BM signature that were downregulated in Rs have been implicated in tumor progression and unfavorable clinical outcomes across multiple cancer types. For instance, UHRF has been shown to promote different types of tumors and contribute to an immune-related risk signature associated with reduced response to ICI in urothelial carcinoma [95,96]. Along these lines, MPRIP has been linked to cancer cell invasion [97,98], while PLEC supports tumor growth and metastasis across different cancer types [99]. In addition, PDGFRB has been associated with adverse clinical outcomes across multiple malignancies, and its inhibition enhances the efficacy of ICI in preclinical models [100,101].

The 13BM discriminated Rs from NRs in the melanoma cohort (AUC = 0.73). This performance is superior to the average performance of previously published gene signatures in melanoma evaluated across melanoma RNA-seq cohorts, including two [43,44] which were used in our study (AUC range of average performances of each signature: 0.38 to 0.70, based on the metanalysis of Coleman et al. [102]). Notably, none of these previously described signatures used a single-cell discovery strategy in which immune cell specific biomarkers were translated to bulk RNA-seq data for model development, as described herein. Importantly, in our case, the 13BM model demonstrated generalizability, following its application in an external, independent, relatively large dataset from a different cancer type: bladder cancer. As expected, model performance in the validation dataset declined (from AUC 0.73 in the melanoma to AUC 0.64 in the bladder cancer dataset), reflecting differences in tumor type and the type of ICIs used across the 2 cohorts. Nevertheless, the model retained significant discriminating ability between R and NR groups, showing superior performance within the low-TMB subgroup (<10 mut/Mb) of the bladder cancer cohort. Specifically, in this subgroup, which is typically associated with poorer outcomes following ICI therapy [103], the model achieved an AUC of 0.75. These findings suggest that the model may help identify responders who fall below current TMB-based stratification thresholds [9]; thereby increasing sensitivity in identifying Rs

The 13BM score in the bulk melanoma tissue demonstrated a strong correlation with transcripts representing immune-related pathways, suggesting that in this context, the signature reflects a “hot tumor” phenotype, typically associated with favorable ICI responses [104]. However, in the bladder cancer cohort, surprisingly, the score had strongest positive correlations with metabolic processes, such as oxidative phosphorylation and mitochondrial activity. Such metabolic pathways have been linked to enhanced T-cell activation within the TME [105]; nevertheless, a better understanding of this result would require a more detailed analysis of the tissue cell components in each tumor type. Interestingly, across both tumor types, the score consistently correlated negatively with ECM organization, EMT, and response to TGF-b. These three processes often function complementarily [106] and are well-documented drivers of immune evasion and immunotherapy resistance [50,107,108]. Collectively, these consistent findings suggest that even though the extraction of the 13BM signature was based on differential expression patterns in immune cells of Rs versus NRs, it also serves as a read-out of not only immune cells but also broader changes of the TME, which eventually can affect ICI response or resistance.

The integrative analysis poses some limitations that should be considered when interpreting the results. As all datasets were publicly available, detailed technical parameters (e.g., library preparation protocols, sequencing depth, or quality control metrics) from the original studies were not always accessible, potentially introducing unaccounted variability. We mitigated this using Harmony integration and the inclusion of each ‘study’ as a covariate for the single-cell data DESeq2 analysis, and with batch correction prior to DESeq2 analysis for the bulk datasets. The pseudobulk approach applied for the analysis, combined with the small cohort sizes, precluded the use of multiple-testing corrections; therefore, nominal p-values were used. Despite this fact, robustness was supported by the consistent replication of the findings across independent datasets, which drastically restrict the possibility of type I errors.

## Conclusion

In conclusion, this study highlights the integral roles of pre-treatment gene expression programs in immune cells in modulating hosts’ responses to ICI. Specifically, a notable association between the T-cell intrinsic expression of the TNFRSF receptor genes and resistance to immunotherapy was observed. The expression of these co-stimulating proteins on the membrane of T-cells may reflect dysfunctional activation and terminal exhaustion, providing a potential biological rationale for the limited efficacy observed in clinical trials testing agonists of these receptors. The single-cell derived expression programs were then distilled into a robust 13BM signature detectable in bulk melanoma but also bladder cancer tissue and predictive of ICI response. Importantly, in patients with low TMB, implementation of this model could help identify Rs, who might otherwise be excluded from treatment based on the approved TMB thresholds. Further validation of the clinical utility of the presented model in parallel to a closer inspection of the spatial expression patterns of the 13 genes in the bulk tissue towards the better understanding of their role in the TME is warranted.

## List of abbreviations

13BM: 13 Biomarkers Model
AUC: Area Under the Curve
CI: Confidence Interval
CR: Complete Remission
CTLA-4: Cytotoxic T-Lymphocyte-Associated Protein 4
DC: Dendritic Cell
DEG: Differentially Expressed Gene
ECM: Extracellular Matrix
EMT: Epithelial-to-Mesenchymal Transition
GO:BP: Gene Ontology Biological Processes
HCC: Hepatocellular Carcinoma
HSC: Hematopoietic Stem Cell
ICI: Immune Checkpoint Inhibitor
IFN-γ: Interferon-gamma
LAG-3: Lymphocyte-Activation Gene 3
MAIT: Mucosal-Associated Invariant T-cell
NRs: Non-Responders
ORA: Overrepresentation Analysis
ORR: Overall Response Rate
OS: Overall Survival
PBMC: Peripheral Blood Mononuclear Cell
PC / PCA: Principal Component / Principal Component Analysis
PD: Progressive Disease
PD-1: Programmed Cell Death Protein 1
PD-L1: Programmed Death-Ligand 1
pDC: Plasmacytoid Dendritic Cell
PR: Partial Remission
Rs: Responders
ROC: Receiver Operating Characteristic
scRNA-seq: Single-cell RNA Sequencing
SD: Stable Disease
SNN: Shared Nearest-Neighbor
TEMRA: Terminally Differentiated Effector Memory T-cells re-expressing CD45RA
TF: Tumor Free
Tfh: Follicular Helper T-cell
TGF-β: Transforming Growth Factor-beta
TMB: Tumor Mutational Burden
TME: Tumor Microenvironment
TNFRSF: Tumor Necrosis Factor Receptor Superfamily
TPM: Transcripts Per Million
Treg: Regulatory T-cell
UMAP: Uniform Manifold Approximation and Projection
UMI: Unique Molecular Identifier

## Declarations

### Ethics approval and consent to participate

Not applicable. This study is using publicly available patient data.

### Consent for publication

Not applicable.

### Availability of data and materials

The datasets analyzed during the current study are available in the Gene Expression Omnibus (GEO), ArrayExpress, and the KU Leuven Research Data Repository. Specifically, single-cell data were retrieved under accessions E-MTAB-13770, GSE120575 and DOI: 10.48804/GSAXBN. The Bulk RNA-seq data were retrieved from GSE78220, GSE213145, E-MTAB-11729 and https://github.com/riazn/bms038_analysis. The code generated in this study is deposited in https://github.com/theomargel/scRNA_Immunotherapy_response.

## Authors’ Contribution

TM: Conceptualization, Data Curation, Data Analysis, Data interpretation, Writing first Draft

IM: Data Analysis, Data interpretation, Manuscript Writing AT: Data collection, Dara interpretation, Manuscript writing

EG: Data analysis, Data interpretation, Manuscript review and editing

SK: Clinical Data analysis, Data interpretation, Manuscript review and editing

MF: Supervision, conceptualization, Funding Acquisition. Data Interpretation, Manuscript Review & Editing.

AV: Supervision, conceptualization, Funding Acquisition. Data Interpretation, Manuscript Review & Editing.

## Data Availability

All data produced in the present work are contained in the manuscript

## Acknowledgements

The authors would like to thank Foteini Paradeisi for her valuable assistance with the creation of the figures. We also acknowledge the support of the MULTIR project consortium.

