## Supplementary File for "Integrative single-cell profiling of melanoma reveals a tumor microenvironment signature predictive of immunotherapy response"

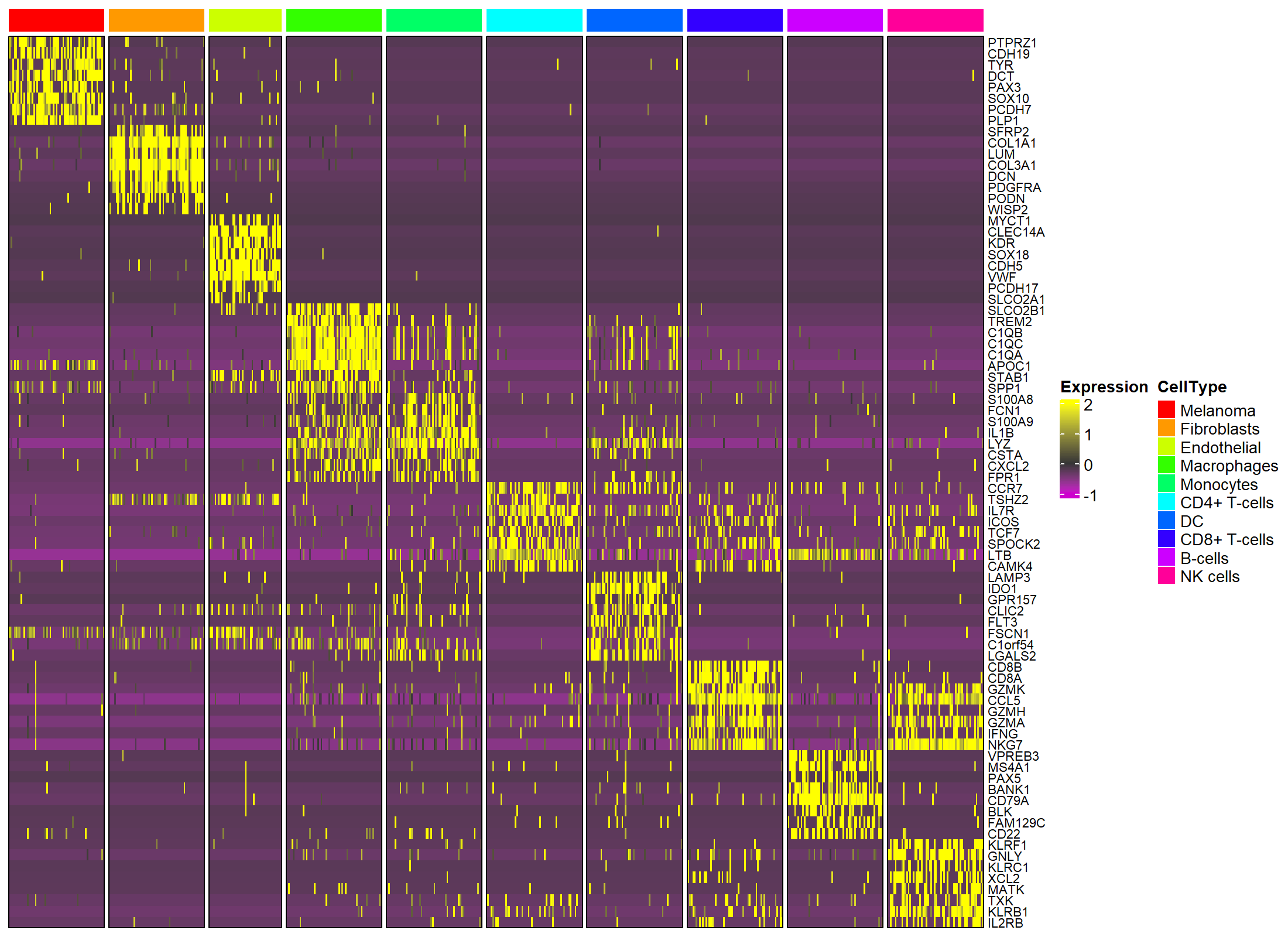

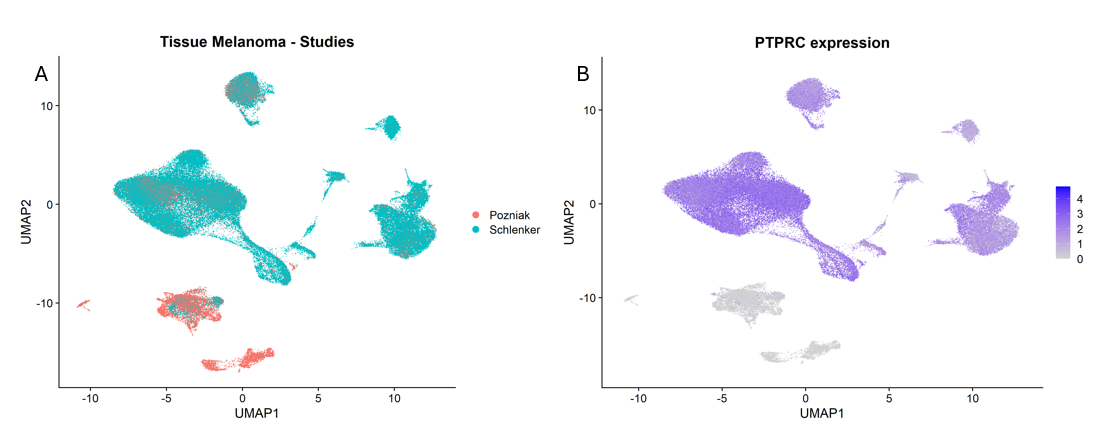
**Supplementary Figure 1.** A) The Studies from which the cells from the discovery single cell cohort derived from, B) The expression of the PTPRC (CD45) immune cells’ marker in the discovery scRNA cohort. It is visible that clusters of cells annotated based on Stromal markers (Figure 2. Original manuscript) do not express PTPRC.

**A**

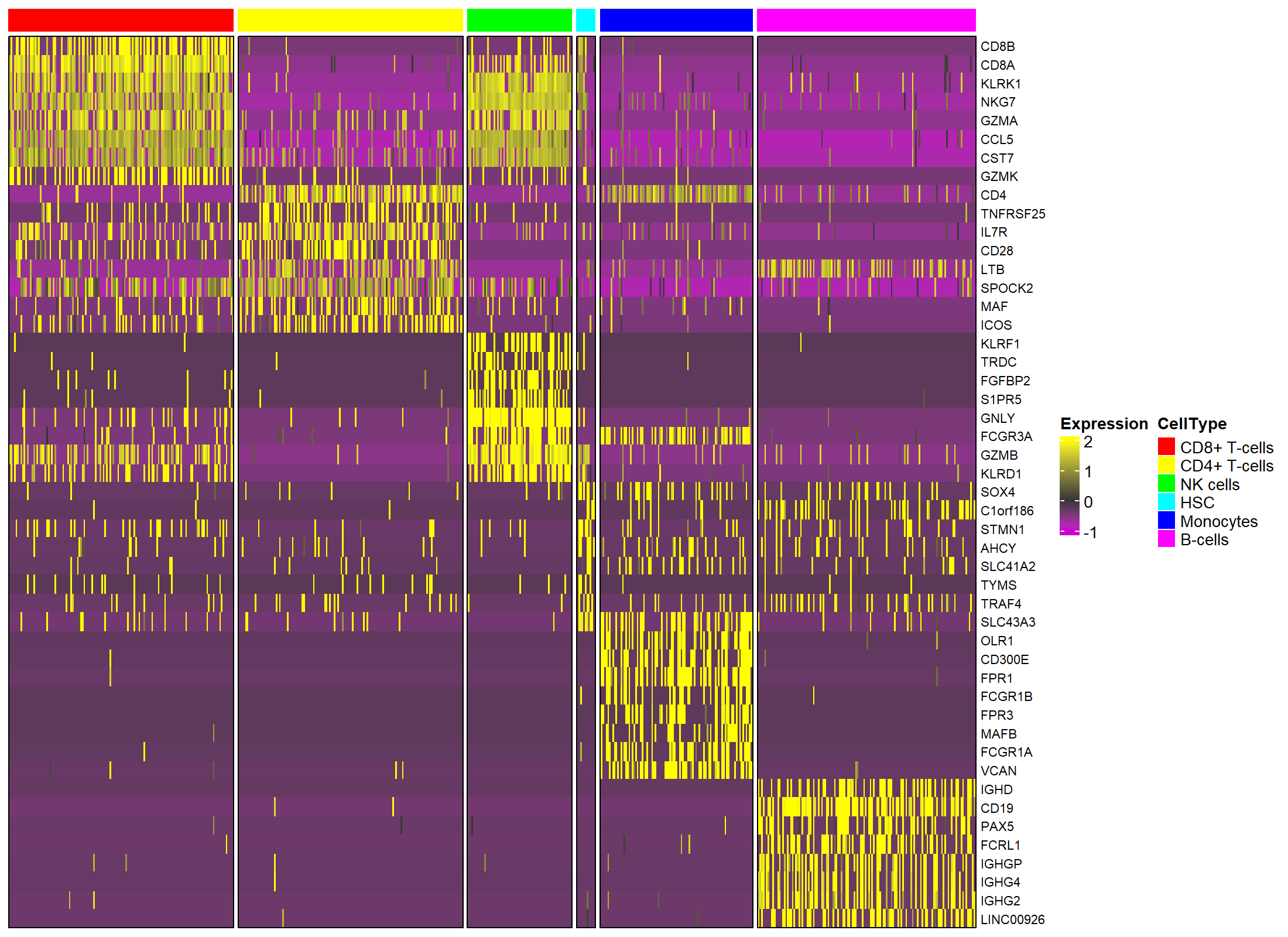
**Supplementary Figure S2.** Transcriptional Validation of Cell Type Identities. Heatmaps displaying the top differentially expressed marker genes across identified cell types in the **(A)** discovery and **(B)** validation datasets. Marker genes were identified using the Seurat FindAllMarkers function. Cell types are indicated by the color bar at the top, and rows represent individual marker genes. Yellow indicates high expression (z-score), while purple indicates low expression.

**B**

**Content of Supplementary Files (In Excel format):**

Supplementary Table 1:

DESeq2 results from the comparison between Rs and NRs in the pseudobulk scRNA-seq discovery cohort. Each spreadsheet tab contains the pseudobulk differential expression results per specific immune cell type (CD4+ T-cells, CD8+ T-cells, NK cells, Monocytes, B cells).

Supplementary Table 2:

Limma results from the comparison between Rs and NRs in the pseudobulk scRNA-seq validation cohort. Each spreadsheet tab contains the pseudobulk differential expression results per specific immune cell type (CD4+ T-cells, CD8+ T-cells, NK cells, Monocytes, B cells).

Supplementary Table 3:

Genes with significant (p < 0.05) expression changes in R versus NR in both  discovery and validation cohorts. Each spreadsheet tab lists the consistently  differentially expressed genes in discovery and validation cohorts per specific immune cell type (CD4+ T-cells, CD8+ T-cells, NK cells, Monocytes, B cells).

Supplementary Table 4:

Table with the Seurat FindAllMarkers output (default settings) showing significantly upregulated genes across T-cell functional states. The analysis is restricted to T-cell-specific genes identified in the original publication (Table 2) and the 13BM model.

Supplementary Table 5:

Results from the differential expression analysis between Rs and NRs in the bulk melanoma integrated dataset. Output from DESeq2.

Supplementary table 6:

Gene set overrepresentation analysis (ORA) results of genes correlating with the 13BM signature score with an absolute value of Spearman |r| > 0.3. The gene set reference was the Gene Ontology (GO) Biological Processes. Sheets named ‘positive correlation-melanoma' and ‘negative correlations-melanoma’  display the pathways enriched in the bulk melanoma cohort, while sheets named ‘positive correlations-bladder cancer' and ‘negative correlations-bladder cancer' contain the pathways enriched in the bulk bladder cohort.
